# In vivo quantification of knee-orthosis valgus corrective moment during gait: Effect of medial strut length on EKAM in medial knee osteoarthritis

**DOI:** 10.1101/2025.11.17.25340432

**Authors:** Kosuke Nakano, Yasuhiro Mine, Junji Katsuhira, Kantaro Yamauchi, Masateru Kitashiro, So Nomoto

## Abstract

Osteoarthritis (OA) is a progressive degenerative articular disease, and knee orthoses are widely used to reduce the external knee adduction moment (EKAM). However, quantitative evidence regarding the corrective moments exerted by orthoses during gait and their relationship with EKAM is scarce. This study directly measured the corrective moments generated by a knee orthosis using a six-axis force sensor and assessed the influence of orthosis structure on these moments and EKAM. Sixteen individuals with medial knee OA (Kellgren–Lawrence grade II or III) affecting the left knee performed walking assessments across three test states: without an orthosis, with an orthosis incorporating a 152-mm medial lower-leg strut, and with an orthosis incorporating a 192-mm strut. Kinematic data and ground reaction forces were used to calculate EKAM, while corrective moments were collected concurrently. The 192-mm orthosis generated greater corrective moments than the 152-mm orthosis throughout all gait phases. EKAM during the loading response declined markedly, being lowest with the 192-mm orthosis, intermediate with the 152-mm orthosis, and highest without an orthosis. An inverse association was observed between corrective moments and EKAM, indicating that higher corrective moments corresponded to lower EKAM. Although walking speed slightly decreased with greater corrective moments, stride length was unchanged. These results demonstrate that the length of the medial lower-leg strut determines the magnitude of corrective moments and is pivotal for lowering EKAM. Collectively, the results highlight the importance of optimizing orthosis design to meet the specific biomechanical requirements of patients with medial knee OA.

## Introduction

The global aging of the population is accelerating, driving a growing incidence of bone and joint disorders associated with degenerative changes. In Japan, approximately 36.23 million individuals—accounting for 29.1% of the total population—are aged 65 years or older, with this share projected to rise further [1]. According to the World Health Organization, the global share of individuals aged 60 years or older is projected to increase from 12% in 2015 to 22% by 2050 [2].

With demographic aging, the prevalence of knee osteoarthritis (OA) continues to increase, affecting approximately 20–38% of individuals aged 65 years or older in countries such as Japan and South Korea [3, 4]. Knee OA substantially impairs quality of life by causing chronic pain, restricted mobility, and difficulty performing daily activities, often leading to loss of independence and psychological distress [5, 6]. In addition to its clinical impact, knee OA imposes a substantial economic burden on healthcare systems, as it represents a leading cause of disability and frequently requires long-term medical management or surgical intervention, including total knee arthroplasty [7, 8]. Consequently, noninvasive treatment approaches—such as exercise, weight management, and orthotic intervention—are gaining prominence for managing chronic knee OA [9, 10].

Knee OA is a multifactorial condition influenced by genetic, biological, and mechanical factors, with abnormal mechanical loading representing the chief driver of its onset and progression [11–13]. As pathology worsens, patients typically exhibit an increased external knee adduction moment (EKAM) and greater mediolateral joint instability [14, 15]. Although conservative treatments, such as exercise and orthotic therapy, are widely recommended due to their noninvasive nature [16], exercise alone is inadequate for lowering joint load, which hastens worsening [17]. Previous investigations have examined the effects of orthoses on EKAM [18–23]; however, most studies have only compared device-on versus off, without quantifying the valgus corrective moment exerted by the device or clarifying its direct relationship with EKAM.

Recent research has investigated various orthotic interventions to treat medial knee OA. Barati et al. [24] contrasted a biaxial ankle–foot orthosis (AFO) with a lateral wedge insole in a randomized crossover trial. Both devices improved pain, enhanced function, and reduced EKAM and its angular impulse, although the AFO produced greater biomechanical improvements, and clinical benefit was modest. Falahatgar et al. [25] compared a lateral wedge with a subtalar strap (without footwear) against a lateral-wedged insole worn in a sandal. Both devices provided prompt analgesic benefit, with no significant difference in EKAM between them, and both improved pain and functional performance after 1 month. Schwarze et al. [26] performed a randomized crossover trial and reported greater load-reducing effects with an AFO than with laterally wedged insoles (approximately 18% vs. 6% reduction in the first EKAM peak and 11% vs. 5% reduction in the knee adduction angular impulse, KAAI). However, patient-reported outcomes showed comparable improvement, indicating no clear clinical superiority. Furthermore, Wang et al. [27] conducted a meta-analysis and found that foot progression angle modifications influence EKAM and KAAI. Toe-in lowers the first EKAM peak (and KAAI) primarily in healthy individuals. Conversely, toe-out reduces the second EKAM peak (and KAAI) in patients with medial knee OA—highlighting the role of gait adaptations in modifying medial knee loading. However, these studies did not measure precisely the corrective moment generated by knee orthoses during gait, leaving it unclear how structural parameters, such as medial strut length, influence corrective forces and their relationship with EKAM.

Against this background, one study proposed a method using two force sensors positioned on the medial lower-leg section of the orthosis—where load is expected to concentrate under the three-point pressure system—to measure the forces generated by the orthosis [28]. Although this method captured localized forces, it did not elucidate the total corrective moment exerted by the orthosis or its overall effect on EKAM. To address this limitation, later investigations examined the relationship between orthosis structure and its corrective forces. For example, one study installed three types of orthoses on a phantom model and attached strain gauges to the structural components to quantify the corrective forces [29]. The results indicated that none of the orthoses produced sufficient corrective force. However, because the evaluation was conducted on a phantom model, the mechanical characteristics under dynamic conditions during actual gait were unassessed.

The magnitude of the valgus corrective moment generated by knee orthoses in ambulation and its influence on EKAM remain unclear. Therefore, experimental studies involving human participants while walking are needed. Furthermore, clarifying the relationship between EKAM and the corrective forces exerted by an orthosis necessitates a system capable of quantitatively assessing the total forces generated by the orthotic structure.

Accordingly, this study sought to quantify the valgus corrective moment generated by a knee orthosis during gait and to examine its relationship with EKAM. Grounded in biomechanics, it was hypothesized that increasing the length of the medial lower-leg strut would enhance the magnitude of the corrective moment. However, individual gait adaptations and alignment variations were likewise anticipated to affect this relationship. To verify these hypotheses, the study measured the actual corrective moment produced by the orthosis, evaluated the effect of medial strut length on the generated moment, and analyzed its association with EKAM in ambulation.

## Materials and methods

This laboratory-based, repeated-measures experimental study took place in the Motion Analysis Laboratory, Toyo University. The study protocol received approval from the Research Ethics Committee of the Graduate School of Life Design, Toyo University (approval Nos. 2023-A14 and 2024-A6S), and complied with the principles of the Declaration of Helsinki. Written informed consent was obtained from all participants before enrollment.

Participants were recruited into this study during two periods: from December 6, 2023, to March 31, 2024, and from July 22, 2024, to March 31, 2025.

Sixteen individuals with left knee OA (seven men and nine women) were enrolled. Missing marker trajectory data caused by occlusion underwent interpolation using the gap-filling function in the Visual3D software.

Baseline demographics, including age, sex, height, weight, affected side, and disease severity based on the Kellgren–Lawrence (KL) classification, are summarized in Table 1. The diagnosis of knee OA underwent medical confirmation following the classification criteria of the American College of Rheumatology [30]. The study included both novice users and habitual wearers of flexible braces. Only participants with KL grades II–III osteoarthritis in the left knee were recruited, representing mild-to-moderate medial compartment OA for which unloading knee orthoses are clinically indicated. Among the participants, nine had KL Grade III and seven had KL Grade II. To minimize variability due to side-to-side asymmetry and ensure standardized testing conditions, we enrolled only individuals with left-sided knee OA.

**Table 1.** Characteristics of patients with knee osteoarthritis (OA)

| Variable | Patients with OA (n = 16) |
| --- | --- |
| Age (years), mean ± SD | 70.1 ± 8.44 |
| Sex (n), female / male | 9 / 7 |
| Height (m), mean ± SD | 160.6 ± 8.04 |
| Weight (kg), mean ± SD | 70.2 ± 14.6 |
| Affected side (n), left/right/bilateral | 16 / 0 / 0 |
| KL Grade (n),<br>left limb (observed), II/III | 7 / 9 |

Abbreviation: SD, standard deviation

Exclusion criteria included inability to ambulate indoors unaided, presence of other orthopedic or central nervous system disorders, communication difficulties, epilepsy, fever, alcohol consumption, or notable pain or swelling on the day of measurement.

The sample size for this pilot study was computed using G*Power 3.1.9.7 (Heinrich Heine University, Düsseldorf, Germany). Assumptions included an effect size of 0.3, statistical power of 0.9, an alpha level of 0.05, a correlation among repeated measures of 0.7, and four measurement sessions. Given these parameters, the minimum required sample size was 14 participants [31–33]. For pilot studies lacking robust preliminary evidence, a minimum of 12 participants per group is generally recommended [34]. Accordingly, the present study met both the statistical and methodological requirements for a pilot investigation.

The data supporting the findings of this study can be obtained from the corresponding author upon reasonable request.

### Experimental setup and protocol

A measurement system based on a center bridge-type knee orthosis (Sakima Prosthetics and Orthotics Co., Ltd., Kunigami District, Okinawa, Japan) assessed the valgus corrective moment generated by the device. The orthosis applies valgus correction through a three-point pressure system, providing support at the thigh and lower leg to reduce EKAM (Fig 1).

**Fig 1.**
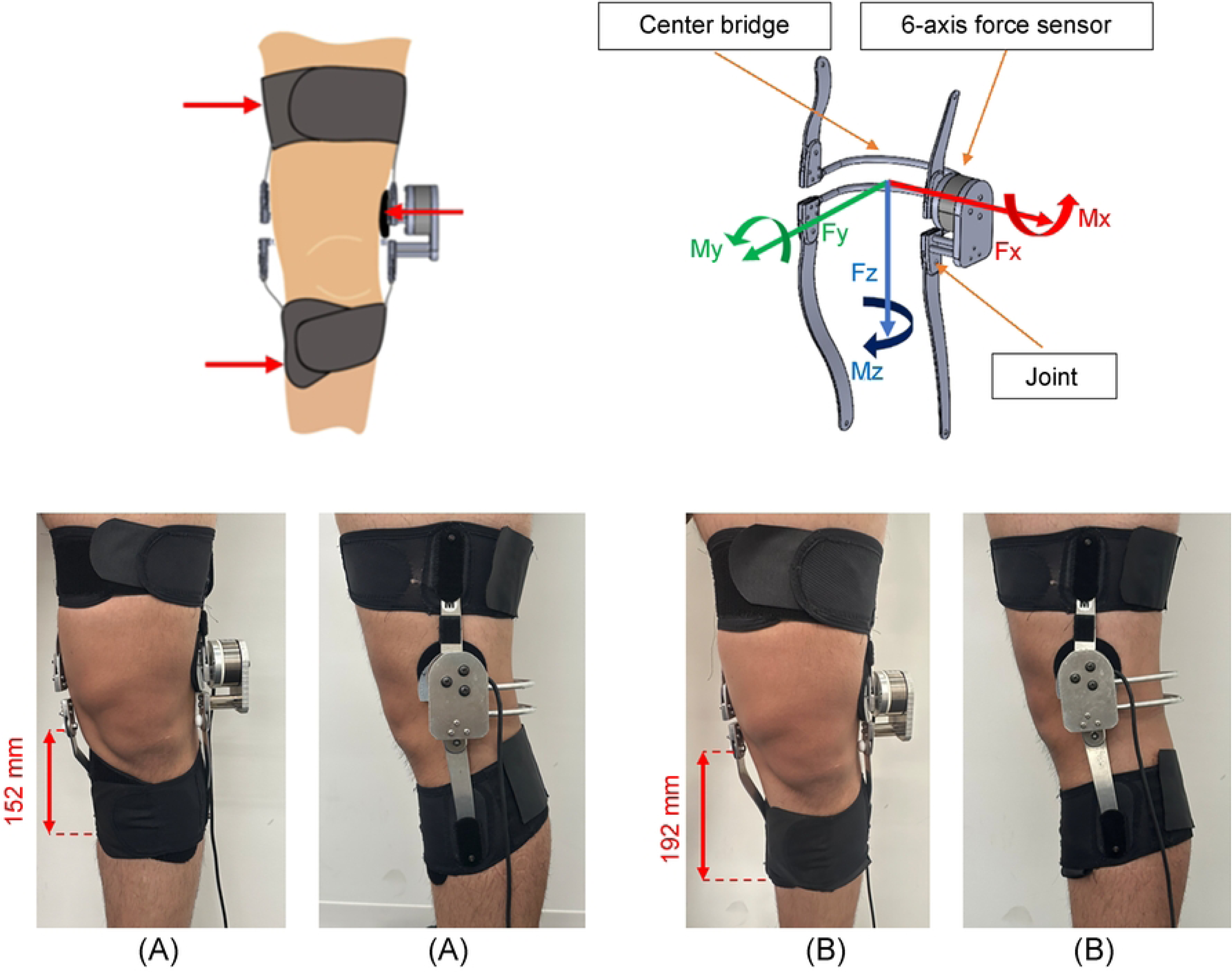
Coordinate framework of the knee orthosis measurement system and orthosis wearing conditions. (A) Knee orthosis with a 152-mm medial lower-leg strut. (B) Knee orthosis featuring a 192-mm medial lower-leg strut A center bridge-type knee orthosis was chosen because its structure provides stable three-point support during gait and enables accurate measurement of the valgus corrective moment. The varus–valgus corrective moment generated between the thigh and shank was quantified by integrating a six-axis force sensor (Leptrino Co., Ltd., Saku City, Nagano, Japan) into the relatively rigid joint section of the orthosis. This sensor simultaneously measures forces and moments along three orthogonal axes, allowing precise acquisition of the relative force and moment data transmitted between the thigh and lower leg.

This configuration permits direct mechanical evaluation of the corrective function of the orthosis, which is difficult to achieve using indirect methods, such as conventional gait analysis. The coordinate framework of the six-axis force sensor used a right-handed convention: the x-axis pointed laterally, the y-axis aligned with the direction of movement, and the z-axis corresponded to the vertical direction (aligned with body weight).

The analysis examined the y-axis moment, representing the varus–valgus corrective moment generated by the orthosis. A negative (−) sign was assigned when the orthosis applied a valgus (abduction) corrective moment to the lower leg relative to the thigh. Conversely, a positive (+) sign indicated a varus (adduction) moment. Gait motion analysis used seven infrared cameras (Vicon MX T series, sampling frequency 960 Hz; Vicon Motion Systems Ltd., Oxford, UK) and three force plates (AMTI, Watertown, MA, USA). These systems enabled the simultaneous, high-precision acquisition of three-dimensional body-segment kinematics and ground reaction forces during walking. Reflective markers (14 mm in diameter) were positioned according to the Helen Hayes protocol at key anatomical landmarks: head (vertex, forehead, and occiput), trunk (seventh cervical vertebra and both acromia), pelvis (anterior and posterior superior iliac spines), upper limbs (elbows and wrists), and lower limbs (thighs, medial and lateral aspects of the knees, shanks, medial and lateral malleoli, and second metatarsals). Additionally, one auxiliary marker was attached to the thigh and shank of the left lower limb fitted with the test knee orthosis, resulting in a total of 34 markers for motion capture (Fig 2).

**Fig 2.**
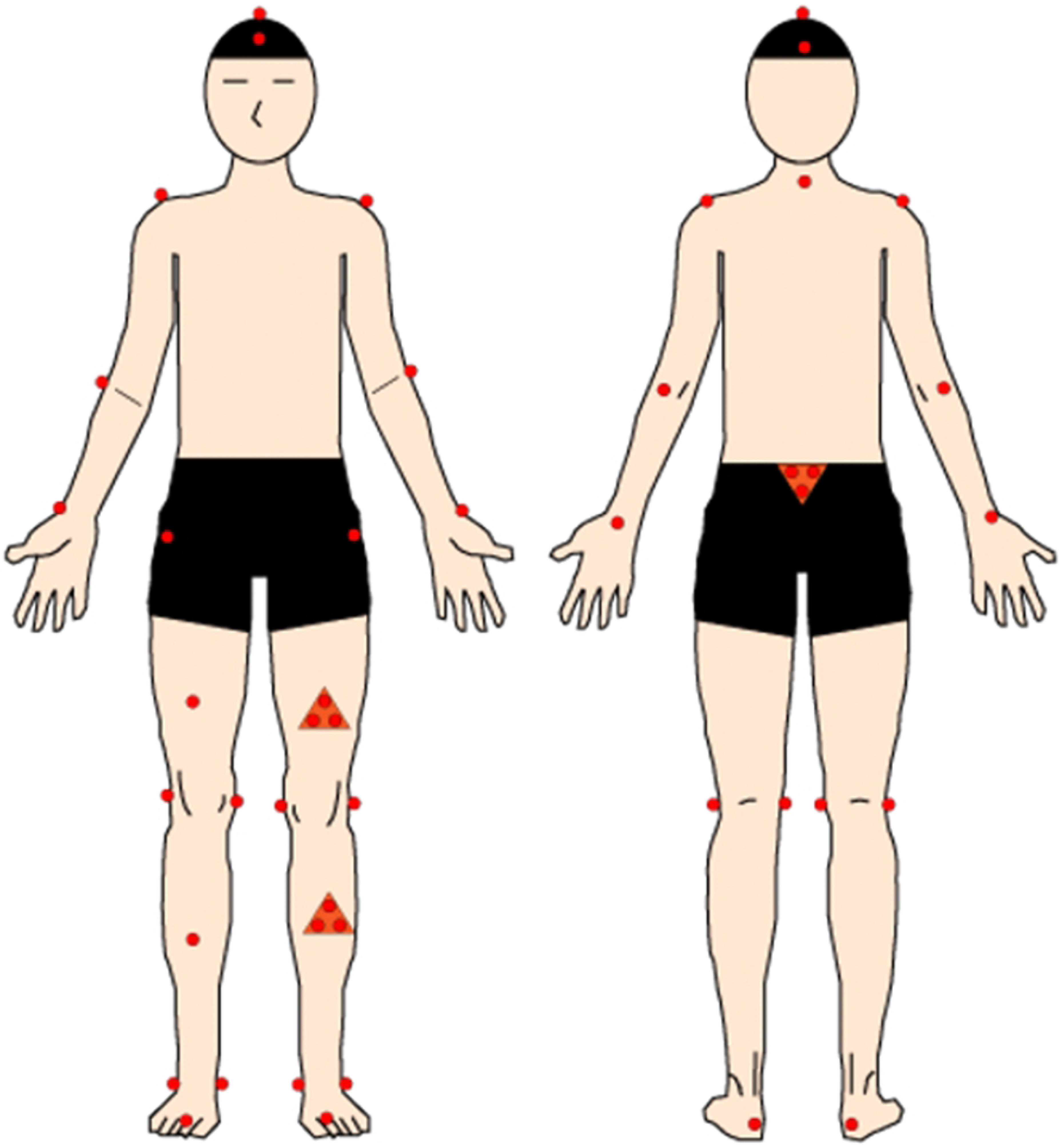
**Locations of reflective markers**. (A) Anterior aspect. (B) Posterior aspect Occasional marker dropouts arose due to occlusion by the knee orthosis or interference from surrounding soft tissue. Missing trajectories were interpolated using the spline function in Vicon Nexus software, with reference to the static standing calibration trial. This procedure ensured both temporal and spatial consistency, thus preserving the robustness and integrity of the dataset. Consequently, no gait trials were excluded from the analysis. Participants completed gait testing under three randomized conditions: (1) without an orthosis, (2) with an orthosis incorporating a 152-mm medial lower-leg strut, and (3) with an orthosis incorporating a 192-mm medial lower-leg strut. The orthotic configurations are depicted in Fig 1. Four gait trials were completed for each condition, yielding a total of 12 trials per participant to ensure adequate data for reliable analysis. Prior to data collection, participants completed a 5-min familiarization period to achieve a natural walking pattern and minimize adaptation effects. Before gait measurements, a 30-s quiet stance posture of the entire body was recorded and used as reference data for marker interpolation and for determining the initial alignment of each body segment. During gait trials, the reflective markers placed on the medial and lateral aspects of the left knee joint were temporarily detached to avoid physical interference with the knee orthosis; their positions were subsequently reconstructed using the static standing data. This interpolation process, conducted in Vicon Nexus software, ensured temporal and spatial consistency throughout the dataset. All gait measurements took place on an 8-m flat, straight walkway. Participants were instructed to walk at a preferred, comfortable speed to maintain natural gait characteristics. During each trial, the valgus corrective moment was sampled continuously using the integrated six-axis force sensor. At the initiation of each measurement, the sensor output was calibrated to zero (0 Nm) under the no-orthosis condition, enabling precise extraction of the corrective moment generated by the orthosis during gait.

## Data analysis

Kinematic and kinetic data were processed in Visual3D software (version 3.6; C-Motion, Germantown, MD, USA). Raw marker data acquired were filtered using a low-pass Butterworth filter with a cutoff frequency of 6 Hz. Similarly, raw data acquired from the ground reaction forces and the Leptrino six-axis force sensor integrated into the knee orthosis measurement system were filtered using a low-pass Butterworth filter with a 15 Hz cutoff frequency. All gait data were time-normalized to span 100% of the stance phase. Knee joint moments and gait performance parameters, including walking speed and stride length, were subsequently computed. Both the knee joint moments and the valgus corrective moments generated by the knee orthosis derived from the six-axis force sensor were scaled to the participants’ body weight.

The analysis focused on the stance phase of the gait cycle, during which considerable mechanical loading occurs at the knee joint. Using the vertical component of the ground reaction force, the stance phase was divided into three subphases: loading response, mid-and terminal stance, and pre-swing. The loading response phase was defined as the period between initial contact of the affected (left) limb and toe-off of the unaffected right lower limb. The mid-and terminal stance phases were defined as the period from toe-off to the next initial contact of the unaffected right lower limb. The pre-swing phase was defined as the period between the initial contact of the unaffected right lower limb and the toe-off of the affected limb.

The integrated values of EKAM and the valgus corrective moment generated by the orthosis were calculated under three test conditions: no orthosis, orthosis with a 152-mm medial lower-leg strut, and orthosis with a 192-mm strut. These values were derived for each subphase—loading response, mid-and terminal stance, and pre-swing—as well as for the entire stance phase. Walking speed and stride length were also analyzed under the same three conditions to assess gait characteristics.

## Statistical analysis

Statistical analyses were performed in JASP software (version 0.19.3.0; JASP Team, Amsterdam, Netherlands). The alpha threshold was set at *p* < 0.05, and all tests were two-tailed. The normality of EKAM, the valgus corrective moment generated by the orthosis, walking speed, and stride length was evaluated using the Shapiro–Wilk test (*p* < 0.05). Parametric tests were used when the assumption of normality was satisfied; otherwise, nonparametric methods were applied. Walking speed and stride length were compared across three test conditions: (1) no orthosis, (2) orthosis with a 152-mm medial lower-leg strut, and (3) orthosis with a 192-mm strut. As the data did not meet the normality assumption, the Friedman test—a nonparametric repeated-measures method—was used. When significant differences were detected, Conover’s post-hoc test was used for pairwise comparisons, with Holm’s correction applied to adjust the familywise error. EKAM was analyzed under the same three conditions using the same procedure. However, because the pre-swing phase of EKAM met the normality assumption, a parametric repeated-measures analysis of variance was used for that phase. Effect sizes were calculated as Kendall’s W for the Friedman tests and as η² for the repeated-measures ANOVA.

According to Cohen’s guidelines, W values of 0.1, 0.3, and 0.5 were interpreted as small, moderate, and large effects, respectively, whereas η² values of 0.01, 0.06, and 0.14 indicated small, moderate, and large effects, respectively [35].

To compare the orthoses with 152-mm and 192-mm medial lower-leg struts, the Wilcoxon signed-rank test—a nonparametric test suitable for paired data—was used. For the Wilcoxon tests, effect sizes were expressed as r values, calculated by dividing the z statistic by the square root of the sample size (r = z/√n). According to Cohen’s benchmarks, |r| = 0.1, 0.3, and 0.5 were interpreted as small, moderate, and large effects, respectively [35]. Finally, Spearman’s rank correlation coefficient was used to evaluate the relationship between the orthosis-generated valgus corrective moment and EKAM under both the 152-mm and 192-mm strut conditions. This nonparametric approach was chosen because it does not assume linearity and is less affected by outliers, making it suitable for small-sample clinical datasets with high variability [36]. The strength of the correlations followed Cohen’s criteria [35]: |r| > 0.1 was considered a weak correlation, |r| > 0.3 a moderate correlation, and |r| > 0.5 a strong correlation.

## Results

### Gait performance

The gait speed and stride length for all orthotic conditions are summarized in Table 2. The mean gait speed without an orthosis was 1.09 ± 0.16 m/s. With the 152-mm medial lower-leg strut orthosis, gait speed was 1.10 ± 0.12 m/s, exhibiting no significant difference compared with either the no-orthosis or the 192-mm orthosis conditions. By contrast, the mean gait speed with the 192-mm orthosis was 1.07 ± 0.13 m/s, which was significantly lower than that with the 152-mm orthosis (*p* < 0.05). Stride length was comparable among the three conditions, with mean values of 1.19 ± 0.08 m (no orthosis), 1.19 ± 0.06 m (152-mm orthosis), and 1.18 ± 0.07 m (192-mm orthosis).

**Table 2.**
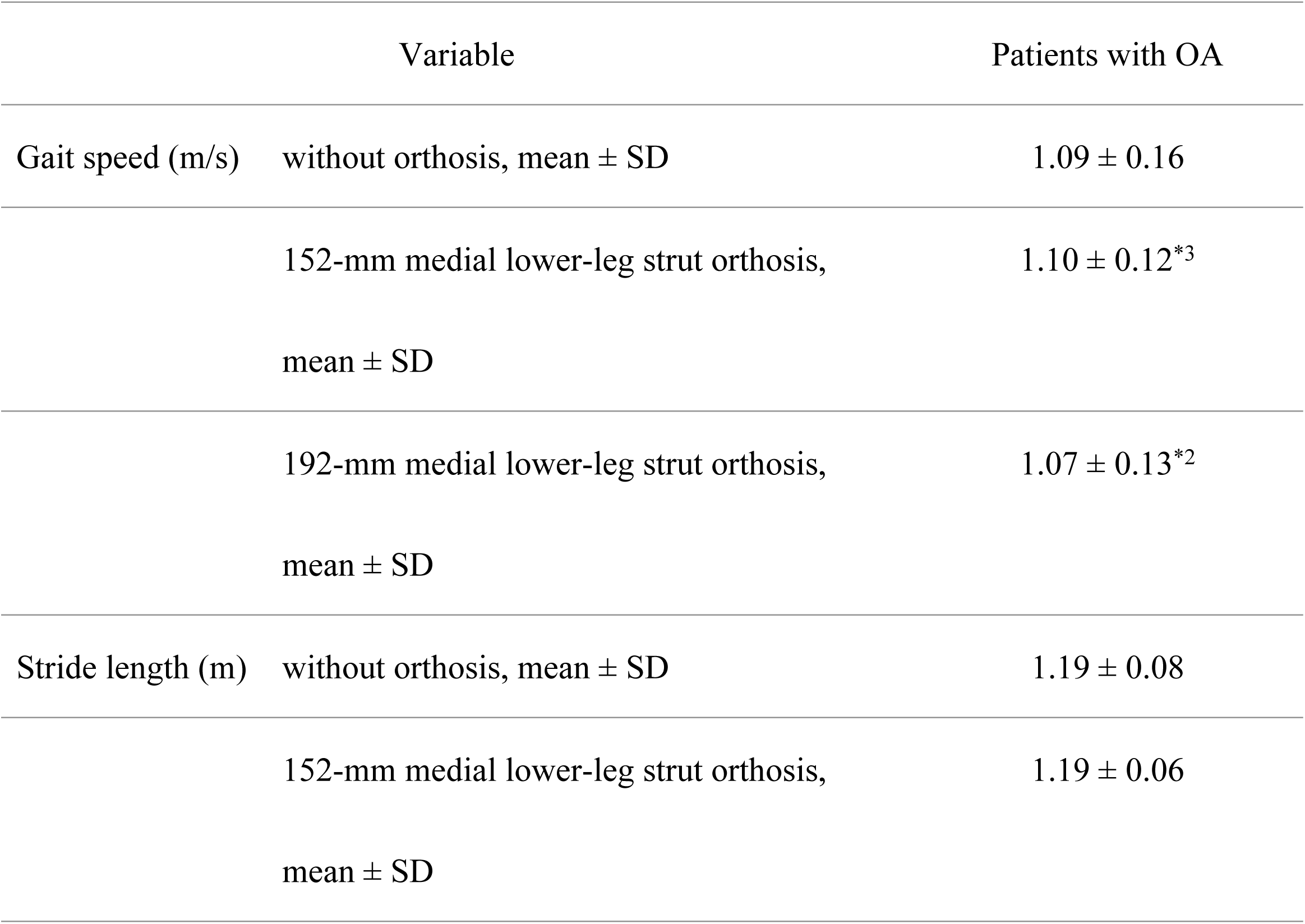
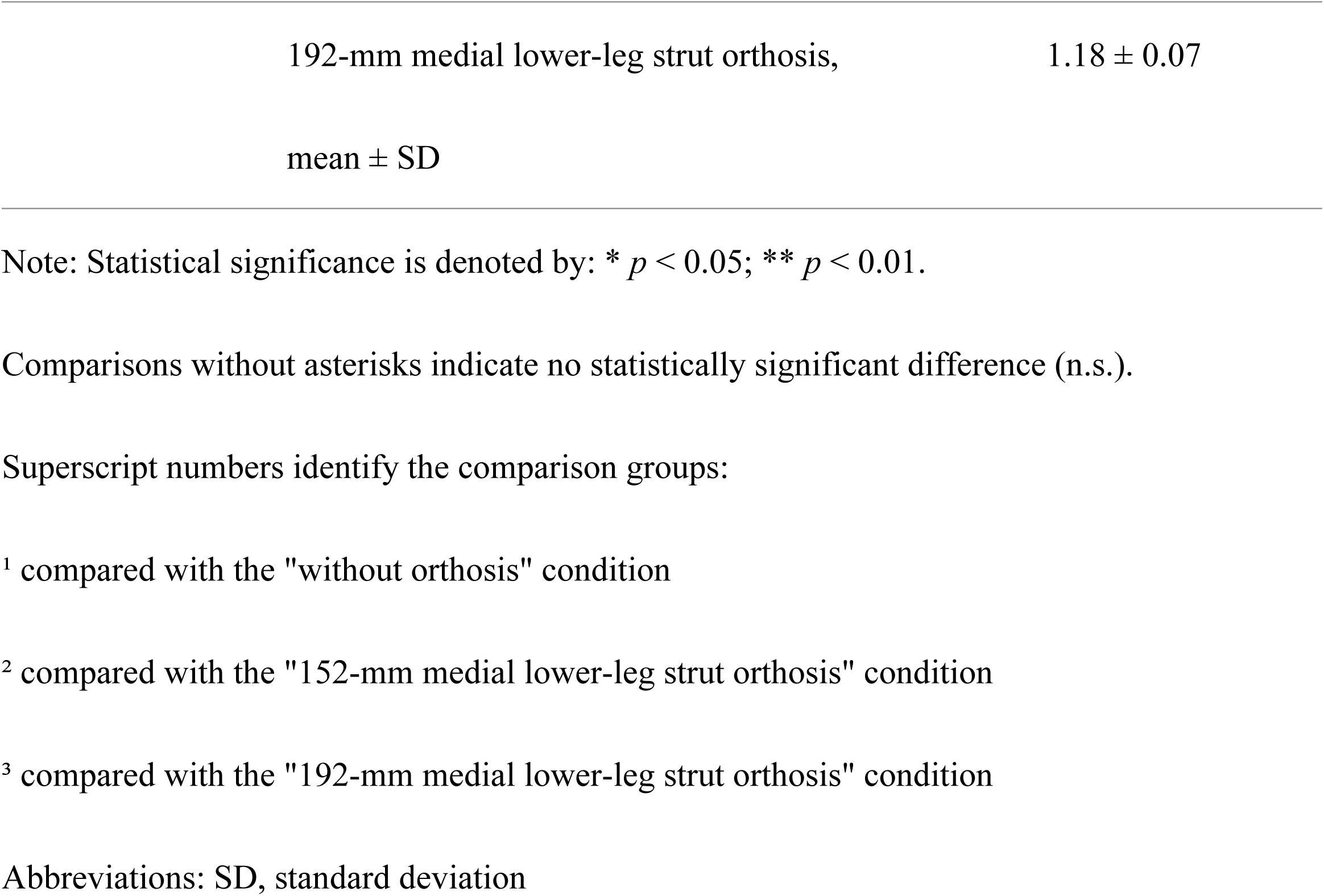
Comparison of gait speed and stride length in patients with knee OA across three conditions: no orthosis, 152-mm strut orthosis, and 192-mm strut orthosis.

### Reduction in EKAM

The EKAM in patients with knee OA was evaluated across the three experimental conditions: no orthosis, 152-mm medial lower-leg strut orthosis, and 192-mm strut orthosis. The results are shown in Table 3 and Fig 3. Under all conditions, a characteristic double-peaked EKAM waveform was observed throughout the stance phase.

**Fig 3.**
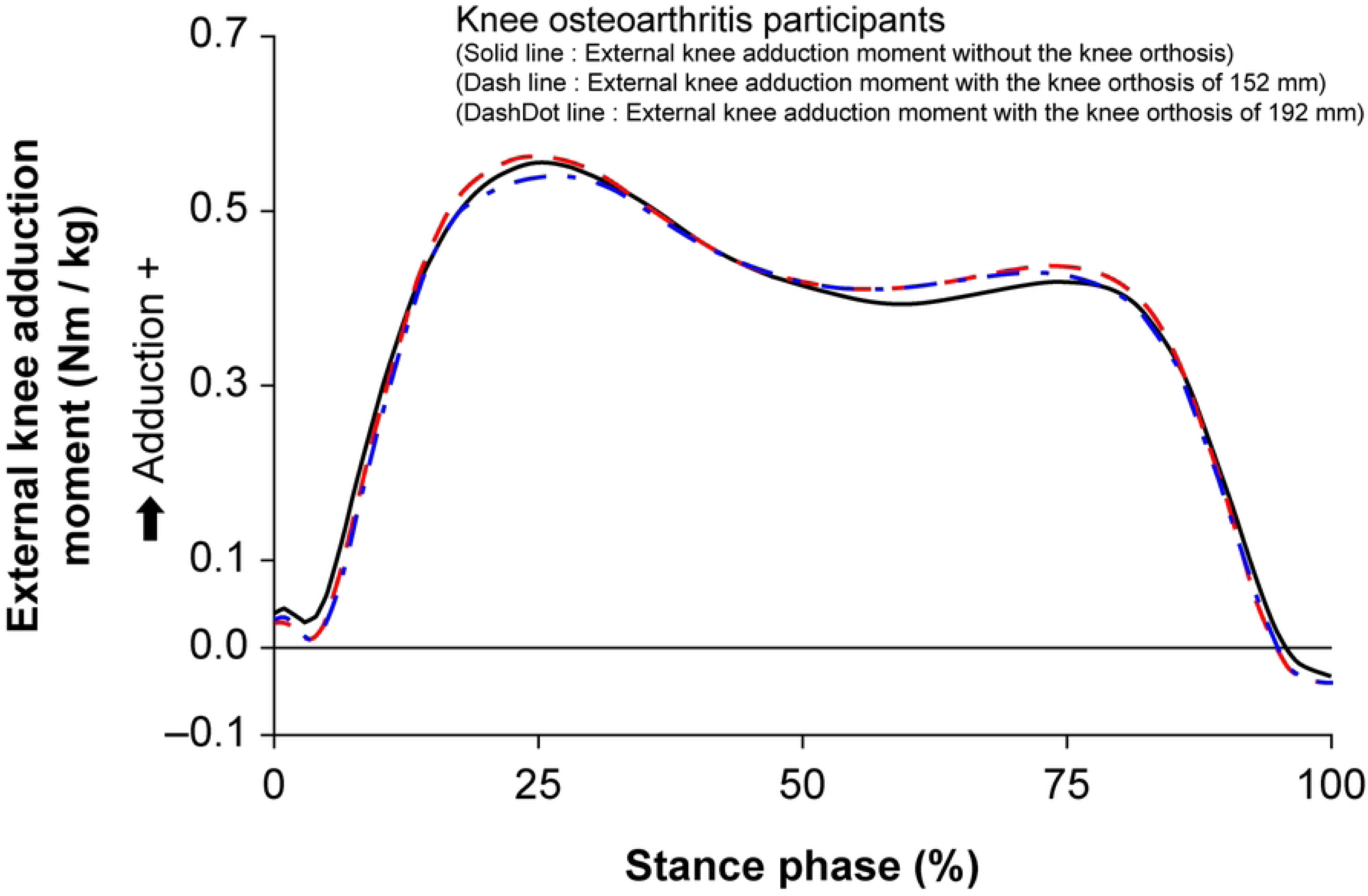
EKAM in ambulation with and without knee orthoses equipped with 152-mm and 192-mm medial lower-leg struts During the loading response phase, EKAM demonstrated a notable reduction when the orthosis was worn. The time-integrated EKAM was 0.022 ± 0.012 Nm·s/kg without an orthosis and significantly decreased to 0.019 ± 0.013 Nm·s/kg with the 152-mm strut (*p* < 0.01). Likewise, with the 192-mm strut, EKAM was further reduced to 0.015 ± 0.011 Nm·s/kg, which was significantly lower than both the no-orthosis (*p* < 0.01) and 152-mm strut (*p* < 0.01) conditions. These reductions are evident in the waveform shown in Fig 3, indicating that both orthosis use and increased strut length contributed to lowering EKAM during the loading response phase. However, during the mid-and terminal stance, pre-swing, and overall stance phases, the waveform patterns were similar across all conditions. Although EKAM during the first peak was markedly reduced with the 192-mm orthosis, the second peak was slightly higher under orthosis conditions, although the difference was minimal. Nevertheless, no statistically significant differences were found among the three conditions during the later phases of stance.

**Table 3.**
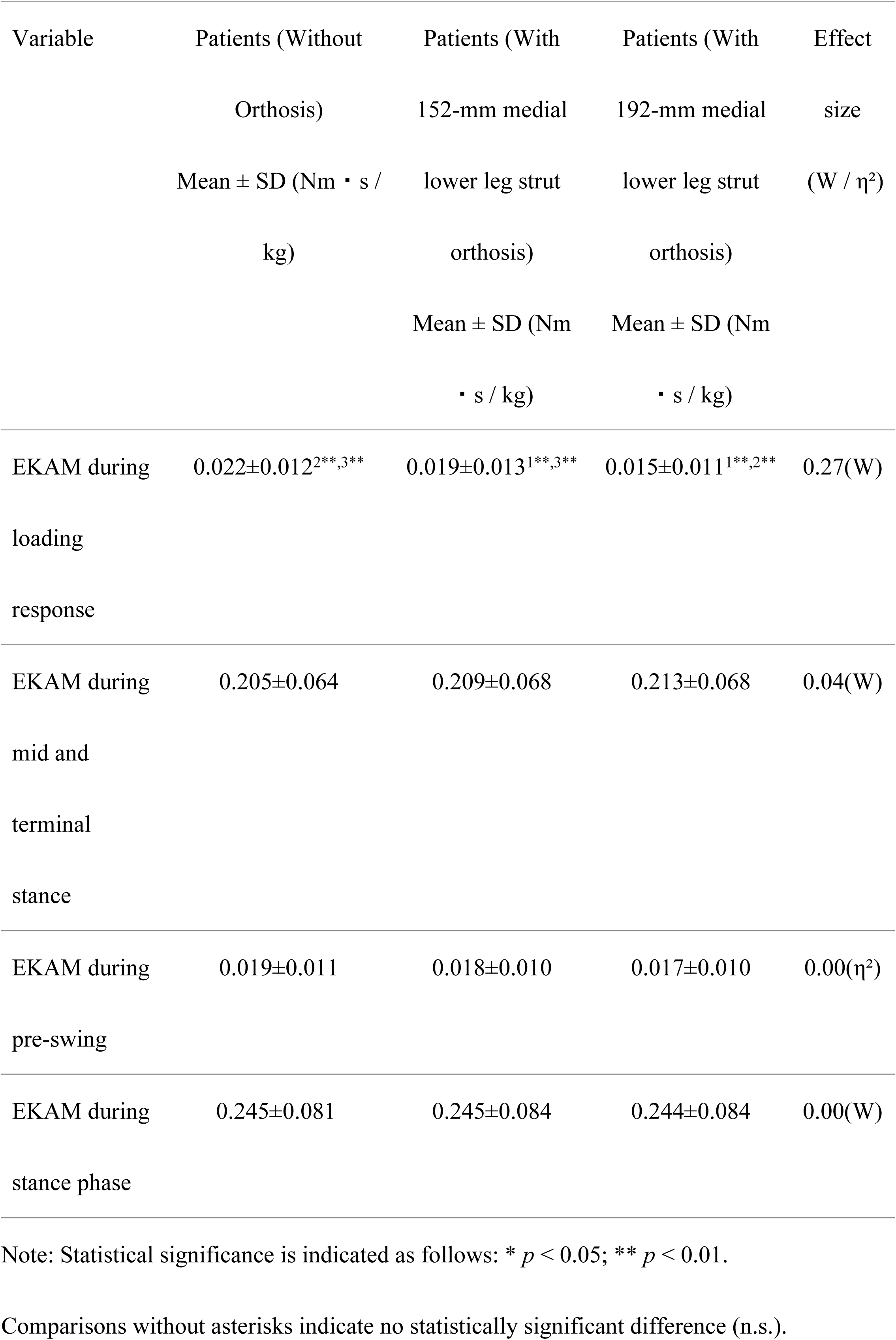
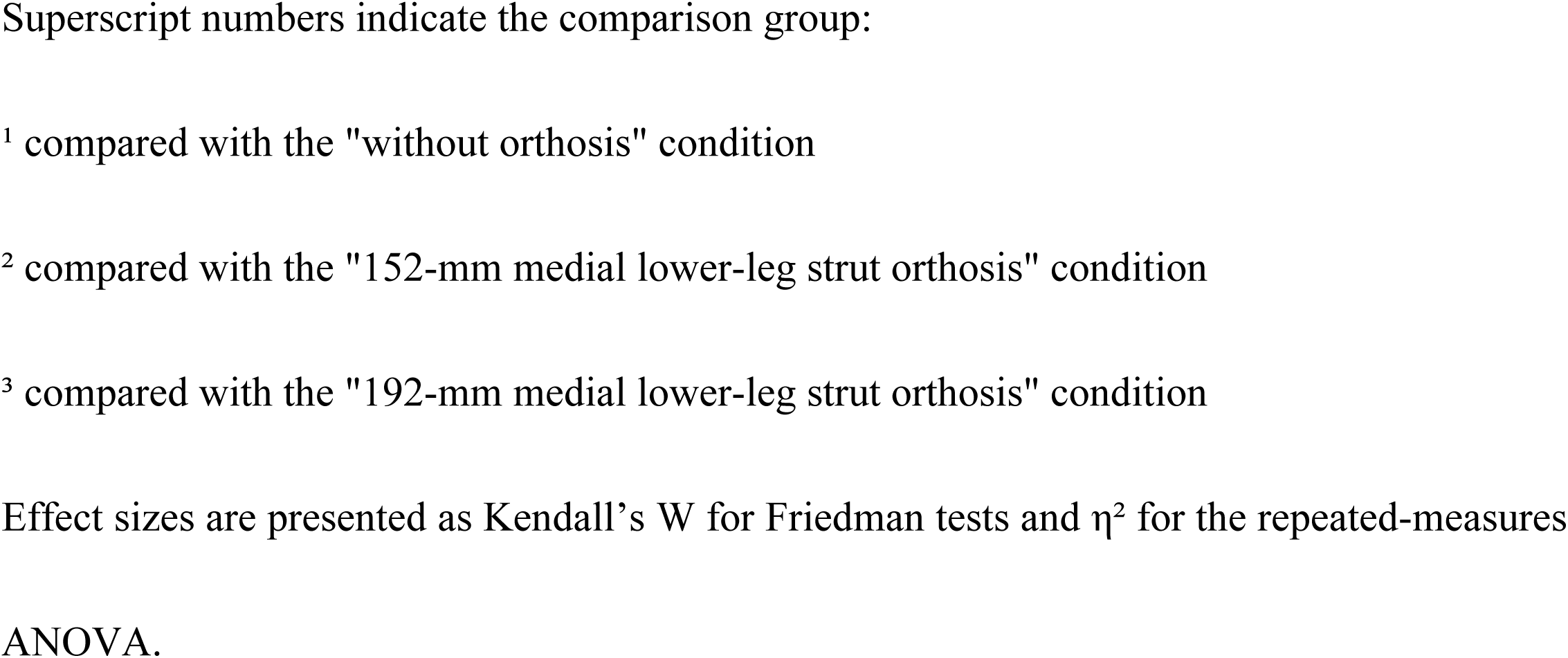
Comparison of the external knee adduction moment (EKAM) in patients with knee OA under no-orthosis and orthoses with different medial lower-leg strut lengths.

The integrated EKAM across the full stance phase was 0.245 ± 0.081 Nm·s/kg without an orthosis, 0.245 ± 0.084 Nm·s/kg with the 152-mm orthosis, and 0.244 ± 0.084 Nm·s/kg with the 192-mm orthosis. No statistically significant differences were detected among the three conditions (*p* > 0.05).

### Valgus corrective moment

The valgus corrective moment generated by the knee orthosis depended on the length of the medial lower-leg strut (Fig 4). Throughout the stance phase, the orthosis with the 192-mm strut consistently generated a greater valgus corrective moment than that with the 152-mm strut. The corrective moment showed a pronounced initial increase during the loading response phase.

**Fig 4.**
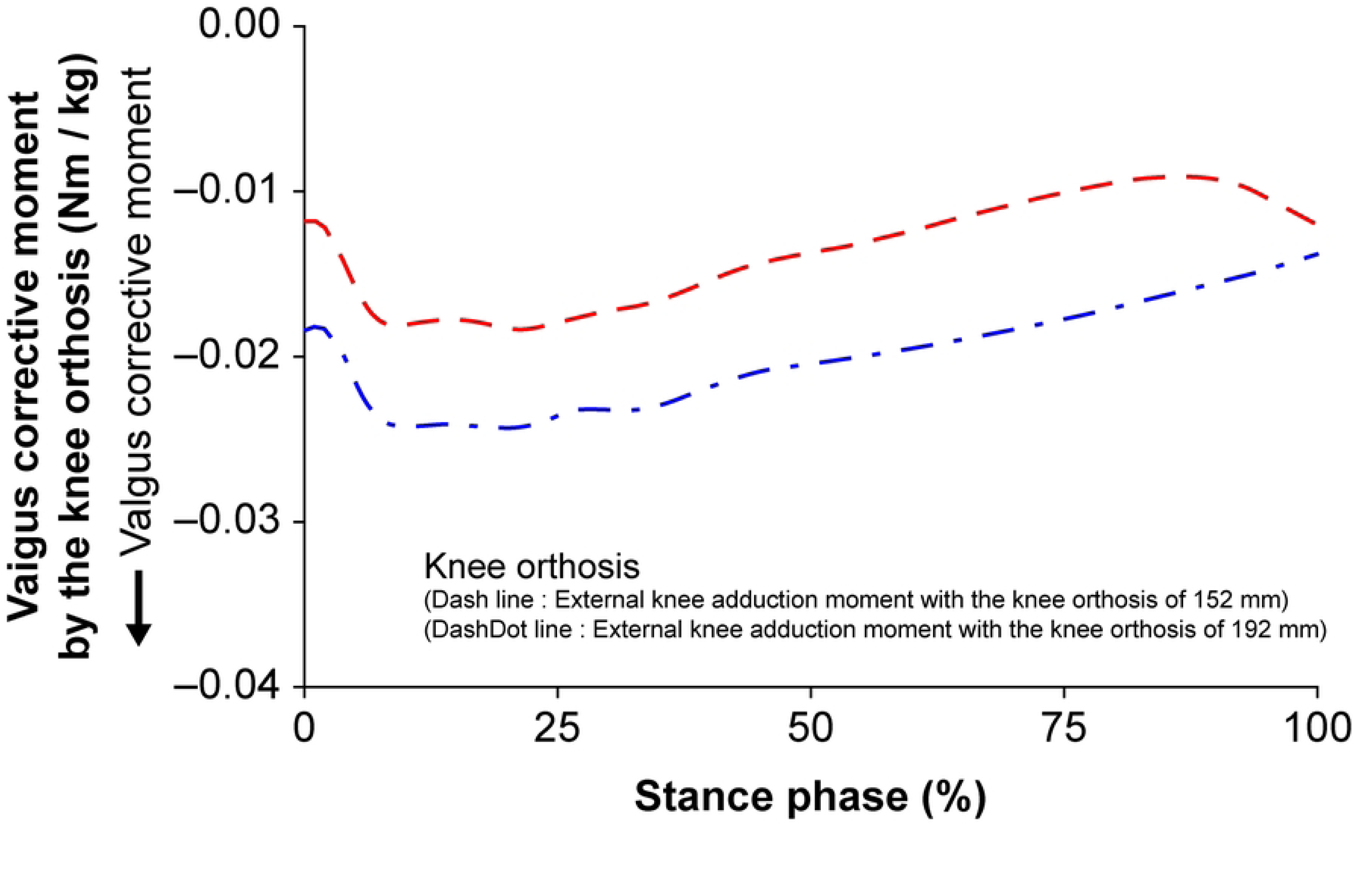
Valgus corrective moment produced by knee orthoses with different medial lower-leg strut lengths Table 4 details the significant differences in corrective moments between the two orthosis configurations. During the loading response phase, the corrective moment was −0.0017 ± 0.00068 Nm·s/kg for the 152-mm strut and −0.0021 ± 0.00089 Nm·s/kg for the 192-mm strut, indicating a statistically significant difference (*p* < 0.01). A similar pattern was observed during the mid-and terminal stance phases, in which the corrective moments were −0.0063 ± 0.0027 Nm·s/kg and −0.0097 ± 0.0036 Nm·s/kg for the 152-mm and 192-mm struts, respectively (*p* < 0.01).

**Table 4.**
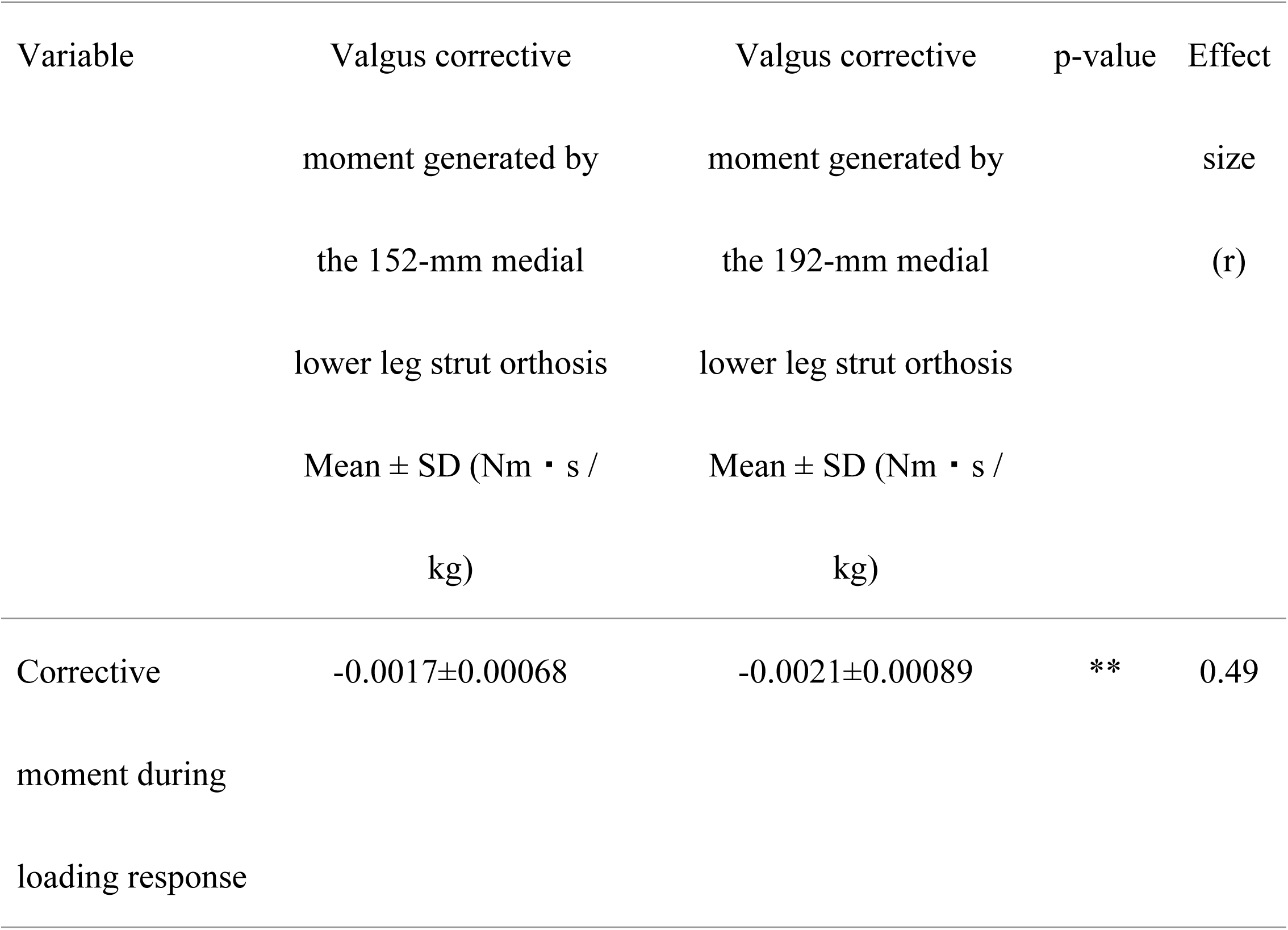
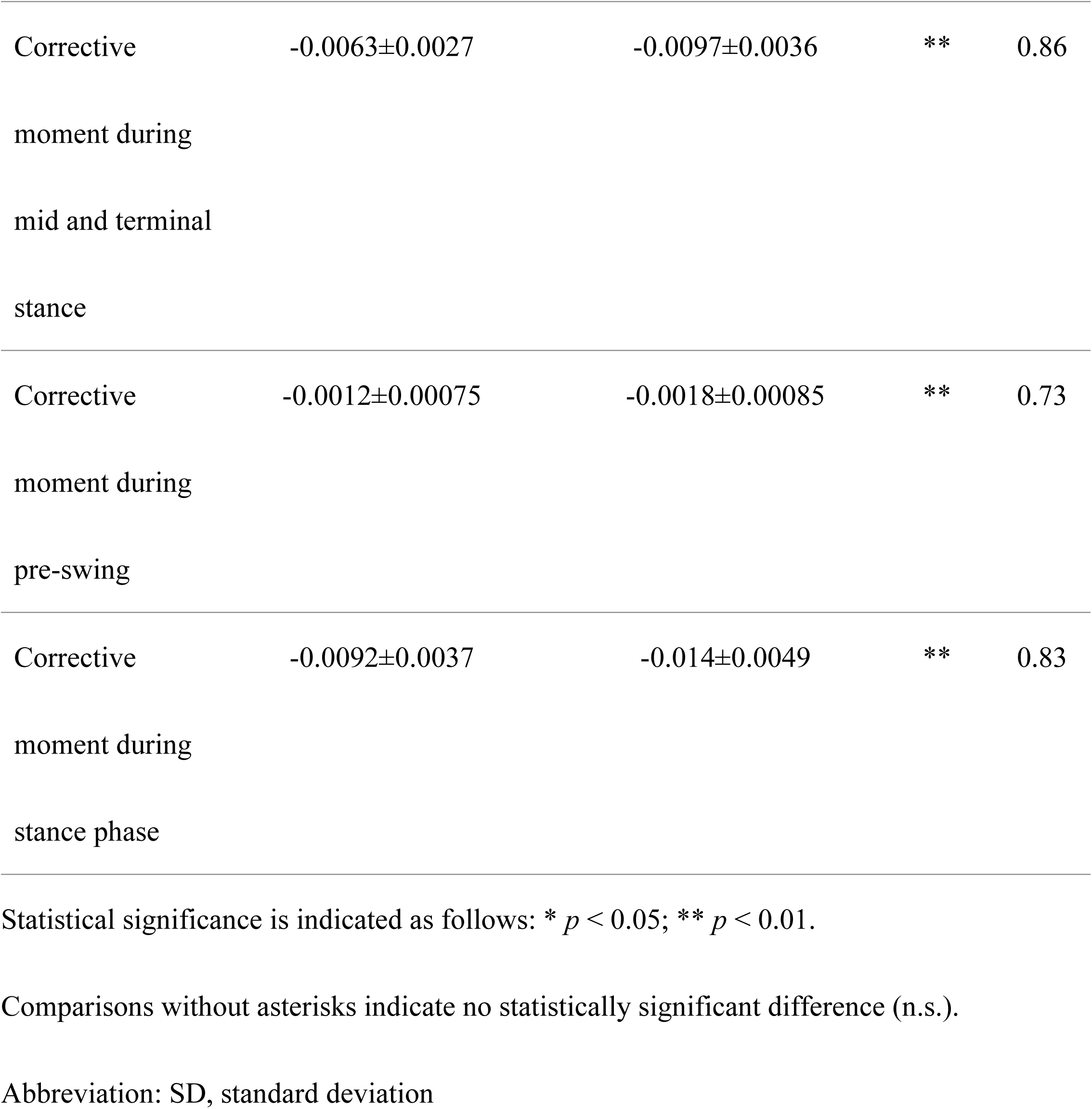
Comparison of valgus corrective moments generated by the 152-mm and 192-mm medial lower-leg strut orthoses.

During both the pre-swing and entire stance phases, the 192-mm strut orthosis consistently showed a significantly larger valgus corrective moment than the 152-mm configuration (*p* < 0.01). Notably, in the pre-swing phase, the corrective moment was −0.0012 ± 0.00075 Nm·s/kg for the 152-mm strut and −0.0018 ± 0.00085 Nm·s/kg for the 192-mm strut. Over the full stance phase, the corresponding values were −0.0092 ± 0.0037 Nm·s/kg and−0.014 ± 0.0049 Nm·s/kg, respectively. Overall, the findings indicate that the 192-mm orthosis generated significantly greater valgus corrective moments across all subphases of the stance phase (*p* < 0.01).

### Relationship between the valgus corrective moment generated by a knee orthosis and EKAM

Fig 5A illustrates the relationship between the valgus corrective moment generated by the knee orthosis and EKAM under the 152-mm medial lower-leg strut condition. A significant negative correlation was observed between the two variables, per Spearman’s rank correlation coefficient (ρ = −0.512, *p* = 0.048). According to Cohen’s (1988) criteria, this correlation constitutes a moderate association between the corrective moment and EKAM. Notably, the coefficient slightly exceeds 0.50, indicating that the relationship nears the cutoff for a strong correlation.

**Fig 5.**
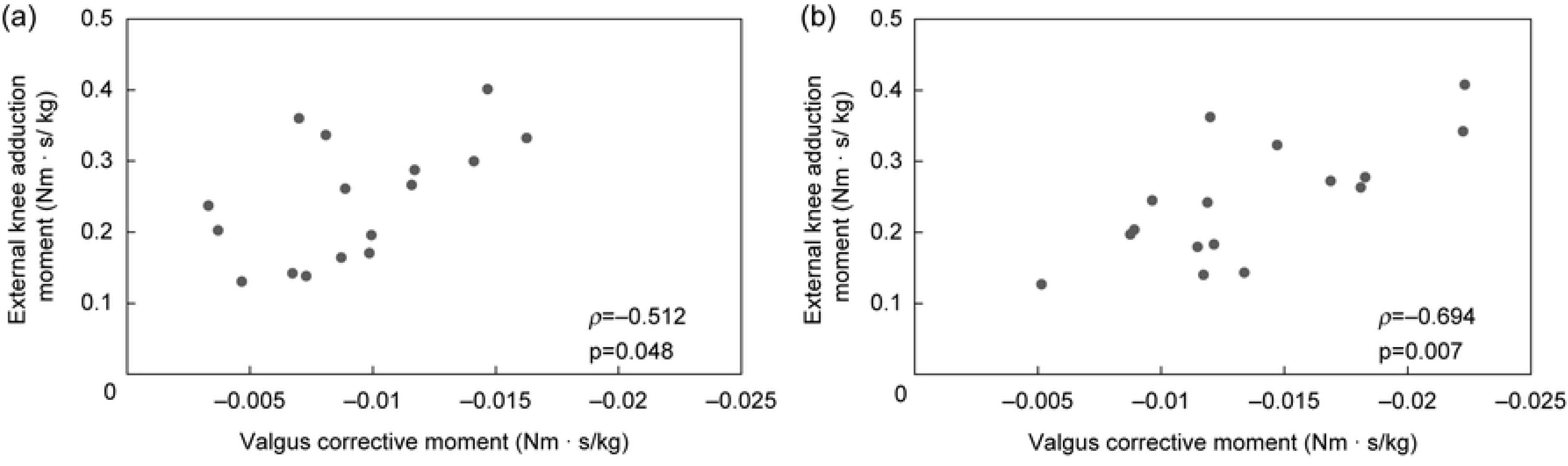
Association between the EKAM and valgus corrective moment generated by the knee orthosis under each condition: (A) 152-mm medial lower-leg strut orthosis. (B) 192-mm medial lower-leg strut orthosis

Fig 5B shows the corresponding relationship under the 192-mm medial lower-leg strut condition, which exhibited a stronger negative correlation (ρ = −0.694, *p* = 0.007). Based on Cohen’s criteria, this correlation qualifies as strong, indicating that the magnitude of EKAM was more closely associated with variations in the corrective moment.

These findings indicate that, under both orthosis configurations, increases in the valgus corrective moment generated by the orthosis were typically associated with decreases in EKAM. The stronger correlation observed with the 192-mm strut suggests that a longer medial lower-leg strut confers more substantial biomechanical effects on EKAM reduction.

## Discussion

This study tested two hypotheses. First, extending the medial lower-leg strut of the knee orthosis would increase the valgus corrective moment, thus lowering EKAM. Second, the relationship between the orthosis-generated corrective moment and EKAM may be influenced by factors such as alterations in lower-limb alignment and gait patterns. Unlike previous studies that merely compared orthosis use versus nonuse, the present study directly quantified the corrective moment using a six-axis force sensor integrated into the orthosis. This method enabled, for the first time, a direct evaluation of the relationship between the measured corrective moment and EKAM during walking. The findings provide novel biomechanical insights into how structural variations in knee orthoses—specifically differences in medial strut length—affect the generated corrective moment and their impact on EKAM.

The first hypothesis was partially supported by the data. Specifically, the 192-mm strut orthosis consistently produced markedly greater valgus corrective moments throughout the stance phase (*p* < 0.01). This increase can be attributed to the extended moment arm from the strut tip to the knee joint center, which theoretically produces a larger moment under an equivalent reaction force. In this study, the 192-mm strut orthosis consistently showed higher corrective moments than the 152-mm orthosis, particularly during the early loading response phase. This finding indicates that orthotic structural parameters play a critical role in load response at initial contact, informing the design of moment-generating mechanisms in knee orthoses.

Similar patterns have been reported in previous studies. Pua et al. [37] demonstrated that even when vertical orthosis dimensions were identical, variations in the relative moment arm—resulting from user height and leg length—influenced orthotic performance. They reported that shorter participants experienced greater reductions in EKAM [37], suggesting that longer relative moment arms increase the generated corrective moment. Collectively, these findings indicate that extending the strut length enhances the production of corrective moments, underscoring the importance of optimizing strut length as a key design factor in clinical orthosis development.

The observed relationship between orthosis structure and EKAM reduction is consistent with previous research showing that valgus knee bracing modifies and typically decreases EKAM during gait [18, 20]. Structural variations, such as differences in orthosis–insole combinations or overall brace configuration, influence both kinematic and kinetic responses [21, 22], thereby shaping the biomechanical effectiveness of the orthosis [23]. Extending these findings from effects to mechanisms, this study directly measured the orthosis valgus-corrective moment, complementing previous comparative analyses of corrective forces across orthosis types [29]. The results show that medial strut length significantly affects both the magnitude of the generated corrective moment and its relationship with EKAM. Furthermore, the minor gait adaptations observed—such as slight reductions in walking speed—accord with previous reports that anthropometric and spatiotemporal factors can influence bracing effects and EKAM magnitude [37, 38].

Our results align with those of Schwarze et al. [26], who reported greater reductions in EKAM with an AFO than with laterally wedged insoles. However, the clinical outcomes of both devices were similar. Similarly, Barati et al. [24] found that both AFOs and lateral wedges reduced EKAM, with AFOs producing greater biomechanical improvements but no commensurate clinical differences. Conversely, Falahatgar et al. [25] observed no significant differences in EKAM between orthotic designs despite symptomatic improvements, underscoring the role of orthosis structure in biomechanical efficacy. Moreover, the gait adaptations observed in this study are consistent with the findings of Wang et al. [27], whose meta-analysis demonstrated that modifications in the foot progression angle can differentially reduce EKAM peaks, showing the interaction between orthosis-generated forces and user-specific gait adaptations.

The second primary objective was to test the hypothesis that the valgus corrective moment generated by the knee orthosis reduces EKAM. The results partly supported this hypothesis. During the loading response phase, both the 152-mm and 192-mm strut orthoses yielded significant reductions in EKAM compared with the no-orthosis condition. Furthermore, the 192-mm strut orthosis achieved a significantly greater EKAM reduction than the 152-mm version. In contrast, EKAM during the mid-and terminal stance phases showed no significant difference between the orthotic and non-orthotic conditions. These findings suggest that the corrective effect of the orthosis is concentrated primarily in the loading response phase, a trend consistent with earlier studies [18, 20], which reported that the influence of knee orthoses tends to be phase-specific within the gait cycle.

The second hypothesis proposed that the relationship between the valgus corrective moment generated by the orthosis and EKAM would be weak owing to potential influences, such as lower-limb alignment and gait pattern changes. However, the results contradicted this expectation: a significant negative correlation was detected between the valgus corrective moment generated by the orthosis and EKAM (*p* < 0.05).

Previous studies have primarily evaluated changes in EKAM based on orthotic presence or absence using inverse dynamic analysis. Consequently, it has remained unclear whether reductions in EKAM result from gait adaptations or from the corrective moment generated by the orthosis itself. By contrast, this study directly measured the corrective moment during gait, enabling quantitative analysis of its relationship with EKAM. A significant negative correlation was found in both cases. For the 152-mm strut, ρ = −0.512 (*p* < 0.05), whereas for the 192-mm strut, ρ = −0.694 (*p* < 0.01), signifying a stronger association with the longer strut. According to Cohen’s criteria, a correlation coefficient of ρ = −0.512 denotes a moderate correlation, whereas ρ = −0.694 indicates a strong correlation. These findings demonstrate a moderate-to-strong negative relationship between the valgus corrective moment generated by the orthosis and EKAM, suggesting that increasing the corrective moment lowers EKAM and that extending the strut length enhances this corrective effect.

Gait speed and stride length were additionally compared across three conditions: no orthosis, a 152-mm strut orthosis, and a 192-mm strut orthosis. No statistically significant differences in stride length were detected, indicating that variations in strut length had minimal influence on step length. However, gait speed was significantly lower for the 192-mm strut than for the 152-mm strut. This reduction likely reflects increased dynamic constraint imposed by the longer strut, which may restrict knee joint mobility and prompt participants to implicitly adjust their gait rhythm. Given that stride length remained unchanged, the observed reduction in gait speed likely stemmed from temporal rather than spatial adjustments.

A similar pattern was reported by Pollo et al. [20], who demonstrated that valgus knee orthosis use significantly lowered EKAM and caused a slight decrease in gait speed. Furthermore, Lusardi et al. [39] noted that both the physical and psychological demands of orthosis use could influence voluntary walking speed, which is consistent with these findings. Additionally, gait speed itself affects EKAM magnitude; Astephen et al. [38] reported that slower walking speeds are often associated with lower EKAM values.

When considered together, these findings suggest that the slight reduction in gait speed observed with the 192-mm strut orthosis may have contributed to the modest decrease in EKAM, alongside the greater valgus corrective moment generated by the orthosis. However, the degree of speed reduction remained within clinically acceptable limits and was unlikely to impair daily activities. From a clinical perspective, optimizing the medial strut length may enhance corrective forces while minimizing gait-related adverse effects, thereby improving both the usability and therapeutic efficacy of knee orthoses for patients with medial knee OA. This study also has educational and practical significance. It is the first to directly quantify valgus corrective moments generated by a knee orthosis during gait with an integrated six-axis force sensor. This approach enhances biomechanical understanding of orthotic function and highlights the role of structural parameters in determining corrective performance. The results further demonstrate that modifying the medial lower-leg strut length alters the generated corrective moment, suggesting that orthotists can tailor orthosis design to individual patient needs. These insights underscore the dual contribution of this research—broadening theoretical knowledge while providing actionable guidance for clinical practice.

This study has several limitations. First, the sample size was relatively small, and potential confounding factors, such as sex, age, and leg length, may not have been fully accounted for. Second, the study examined only the immediate biomechanical effects of short-term orthosis use, without assessing long-term adaptations or patient-reported outcomes. Third, although a significant negative correlation was observed between EKAM and the valgus corrective moment, causality cannot be inferred because alignment changes and gait modifications may also have contributed. Fourth, only medial strut length was assessed; other design parameters, such as material stiffness, fixation method, and user comfort, were not evaluated.

Future research should address these limitations by (i) including larger, stratified cohorts, (ii) evaluating long-term biomechanical and clinical effects, (iii) incorporating complementary analyses, such as intra-articular pressure estimation, electromyography, and soft-tissue stress analysis, and (iv) systematically investigating multiple structural design parameters to develop evidence-based guidelines for orthotic design and clinical application.

## Conclusions

This study examined the effects of medial lower-leg strut length on the valgus corrective moment generated by the orthoses and EKAM in ambulation. The 192-mm strut orthosis produced markedly larger valgus corrective moments than the 152-mm strut across the entire stance phase. This finding suggests that extending the moment arm from the distal end of the strut to the knee joint center increases the generated moment given equal loading. Furthermore, a significant negative correlation emerged between the valgus corrective moment generated by the orthosis and EKAM, particularly under the 192-mm strut condition, where a strong correlation was identified (ρ = −0.694, *p* < 0.01). These results provide quantitative evidence that orthotic structure plays a meaningful role in reducing EKAM. Although stride length did not differ significantly among the conditions, gait speed was slightly reduced with the 192-mm strut orthosis (*p* < 0.05), likely reflecting increased dynamic constraint from the longer strut, which may have prompted implicit cadence changes. In summary, the medial lower-leg strut length materially affected both the valgus corrective moment and EKAM reduction, especially during the loading response phase. These findings highlight medial strut length as a key design factor and inform the creation of biomechanically optimized and individually tailored orthotic strategies for clinical application. This study offers an empirical foundation for future orthosis design and evaluation. Future studies should examine additional design variables—such as overall orthosis stiffness, strap positioning, and material composition—to further optimize the corrective force generated from the device. Longitudinal research is necessary to assess the long-term clinical effectiveness of orthotic use. Moreover, studies involving larger and more diverse populations, encompassing varying severities of knee OA and limb deformities, will be essential for validating and generalizing these findings and advancing personalized orthotic solutions.

## Data Availability

Due to ethical restrictions related to participant privacy, the individual-level raw data cannot be shared publicly. Only anonymized, aggregated statistical data that do not allow the identification of any individual participant are available. These aggregated data are included within the manuscript and its Supporting Information files, in accordance with the approved ethics protocol.

## Acknowledgments

The authors are grateful to Tamotsu Sakima (Chairman) and Ichiro Sakima (President) of Sakima Prosthetics and Orthotics Co. for providing the materials used in developing the knee orthosis measurement system applied herein.

